# A Multi-center, Prospective, Observational-cohort controlled study of Clinical Outcomes following COVID-19 Convalescent plasma therapy in hospitalized COVID-19 patients

**DOI:** 10.1101/2021.06.14.21258910

**Authors:** Lakshmi Chauhan, Jack Pattee, Joshay Ford, Chris Thomas, Kelsey Lesteberg, Eric Richards, Michele Loi, Larry Dumont, Kyle Annen, Mary Berg, Mercedes Zirbes, Amanda Miller, Timothy C. Jenkins, Tellen D. Bennett, Daniel Monkowski, Rebecca S. Boxer, J. David Beckham

## Abstract

**Background:** The SARS-CoV2 pandemic has caused high inpatient mortality and morbidity throughout the world. COVID19 convalescent plasma has been utilized as a potential therapy for patients hospitalized with COVID19 pneumonia. This study evaluated the outcomes of hospitalized COVID19 patients treated with COVID19 convalescent plasma in a prospective, observational multicenter trial.

**Methods:** From April 2020 through August 2020, hospitalized COVID19 patients at 16 participating hospitals in Colorado were enrolled and treated with COVID19 convalescent plasma (CCP) and compared to hospitalized patients with COVID19 who were not treated with convalescent plasma. Plasma antibody levels were determined following the trial given that antibody tests were not approved at the initiation of the trial. CCP-treated and untreated COVID19 hospitalized patients were matched using propensity scores followed by analysis for length of hospitalization and inpatient mortality.

**Results:** 542 total hospitalized COVID19 patients were enrolled at 16 hospitals across the region. A total of 468 hospitalized COVID19 patients were entered into propensity score matching with 188 patients matched for analysis in the CCP-treatment and control arms. Fine-Gray models revealed increased length of hospital stay in CCP-treated patients and no change in inpatient mortality compared to controls. In subgroup analysis of CCP-treated patients within 7 days of admission, there was no difference in length of hospitalization and inpatient mortality.

**Conclusions:** These data show that treatment of hospitalized COVID19 patients with CCP did not significantly improve patient hospitalization length of stay or inpatient mortality.

## Background

The ongoing pandemic due to SARS-CoV2 has caused over 150 million cases and 3 million deaths worldwide as of April 2021. Since the onset of the pandemic, various therapeutic measures including antivirals, immunomodulatory therapeutics, and passive antibody therapies have been evaluated for efficacy in the treatment of COVID19. Convalescent plasma therapy has been utilized for several viral and non-viral infectious diseases throughout history and was the only means of treating certain infectious diseases prior to the development of antimicrobial-specific therapy in the 1940s.[1, 2] Experience from prior outbreaks with other coronaviruses, such as SARS-CoV, shows that convalescent plasma contains neutralizing antibodies to the relevant virus and neutralizing antibodies may provide therapeutic benefit to acutely infected patients.[3] In the case of SARS-CoV2, passive antibody therapy with convalescent plasma may mediate protection by viral neutralization or other mechanisms such as enhanced phagocytosis and antigen processing of virions.

Convalescent plasma, hyper-immune globulin, and monoclonal antibodies are different passive antibody therapeutics evaluated as possible treatments for COVID-19. Recent studies of COVID19 convalescent plasma (CCP) therapy in several studies have shown potential benefit of treatment in hospitalized COVID19 patients.[4-7] Other studies including randomized, open-label trials have exhibited lack of a clear clinical benefit from CCP therapy in hospitalized COVID-19 patients.[8-10] Because, each study exhibits a range of strengths and weaknesses in study design, different targeted patient populations, and study power, additional studies of CCP therapy in hospitalized COVID-19 patients are needed to inform appropriate use of CCP as a potential therapeutic alternative for COVID19.

Our study utilized a multi-center trial design across 16 academic and non-academic hospitals in Colorado in an open-label, prospective, observational, cohort-controlled trial to evaluate length of hospitalization and inpatient mortality rate in COVID-19 patients treated with CCP. We found that CCP treatment of hospitalized COVID19 patients did not significantly alter length of hospitalization or inpatient mortality rates. These data support recent findings showing that treatment with CCP for hospitalized COVID19 patients does not significantly improve clinical outcomes.

## Methods

This was an open-label, prospective, multi-center cohort trial comparing hospitalized patients with COVID-19 who received CCP to hospitalized COVID-19 patients who received standard-of-care treatment. The clinical study was conducted at 16 hospitals in Colorado including UCHealth Metro Denver, UCHealth North, UCHealth South hospitals, Denver Health Medical Center, Children’s Hospital of Colorado, and SCL Health Hospitals. All human clinical trial work on COVID-19 was approved by the Colorado Multi-Institution Review Board (COMIRB, #20-0986, #20-0990) prior to the study opening. All hospitals within networks of UCHealth, Children’s Hospital Colorado, and Denver Health Medical Center provided CCP through a University of Colorado Expanded Access program (FDA IND#21426). SCL Health Hospitals provided CCP through FDA Expanded Access program sponsored by Mayo Clinic. All patients or designated decision makers provided signed informed consent prior to enrollment into expanded access programs to receive CCP. The study was conducted from April 2020 through August 2020.

Each site prospectively screened eligible patients for inclusion based on pre-determined inclusion criteria: age 18 years or older, laboratory confirmed diagnosis of SARS-CoV-2 infection by detection of nucleic acid from a respiratory sample, admission to a participating facility, COVID-19 pulmonary disease requiring hospitalization, sufficient COVID19 convalescent plasma available for treatment, and informed consent provided by the patient or healthcare proxy. Exclusion criteria for the observational clinical trial included receipt of pooled immunoglobulin in the past 30 days, patients placed on extracorporeal membrane oxygenation, history of transfusion reaction, contra-indication to receiving plasma products, and risk of transfusion exceeds potential benefit based on clinician determination.

Control patients were identified, and data captured over each month of the study using a UCHealth database of hospitalized COVID19 patients for each month from April through August of 2020. Data was abstracted from all hospitalized COVID-19 patients in the UCHealth system that met the inclusion and exclusion criteria above but did not receive CCP therapy over the same time period. As previously described, all patient data was entered and maintained in a secure, HIPAA-compliant REDCap database.[11]

### Convalescent Plasma procurement and transfusion

FDA approval of the expanded access program for the use of convalescent plasma was obtained (IND#21426). Convalescent plasma was allocated from FDA-registered blood establishments (Children’s Hospital Colorado, Garth Englund Blood Center, Vitalant Center) to the treating hospitals using standardized procedures. COVID-19 convalescent plasma was supplied as an investigational blood product and was administered according to standard hospital procedures for plasma administration. Plasma was infused over 1-2 hours (rate of 100-250 ml/hr). Pre-medications, such as acetaminophen or diphenhydramine were provided as indicated by the treating physician. One unit of COVID-19 convalescent plasma was administered to anyone weighing less than 90 kg and 2 units were given to patients over 90kg. Levels of binding antibodies were assessed using VITROS anti-SARS CoV2 IgG assay. Some CCP units were provided prior to availability of antibody testing and frozen samples were each retrospectively tested for antibody. Of 375 units of CCP utilized in the study, 362 met FDA criteria for positive CCP antibody > 12. Three tested units were negative for antibody and 10 units were not tested due to lack of frozen sample. The patients that received these units were included in the full intention-to-treat analysis. Ten patients enrolled through SCL Health had CCP units tested through the Mayo Clinic Expanded access program and data was not available for analysis.

### Propensity Score Matching

Propensity score matching was performed to ensure that potential confounding factors were balanced between the CCP treatment group and the control group. Propensity score matching is an analysis approach utilized for non-randomized trials to minimize bias in estimating the treatment effect.[12, 13] Propensity scores are calculated based on baseline criteria at admission that are expected to influence outcome. For example, age over 70 years is known to increase risk of a negative outcome for patients hospitalized with COVID19.[14] Thus, matching patients based on age in the treatment and control groups will help to decrease possible bias of enrolling patients of more advanced age in one group. The propensity score was estimated via logistic regression with no higher order or interaction terms. We conducted greedy nearest neighbor one-to-one propensity score matching via the R package MatchIt.[15] A caliper length of 0.2 multiplied by the standard deviation of the logit of the propensity score was used.[16] Covariate mean balance was assessed via standardized mean difference, with balance defined to be standardized mean difference < 0.1.[17] The matching criteria included ethnicity, age (categorical by decade), admission month (continuous), sex, hypertension, lung disease, cancer, diabetes, obesity, smoking status, and immunosuppression. Lung disease was defined as an underlying lung disorder that requires treatment including asthma, chronic obstructive pulmonary disease, and interstitial lung disease. Immune suppression was defined as any condition that results in suppression of immune responses including primary and secondary immune deficiencies that result from treatment.

### Statistical analysis

We modeled the primary outcome (time to hospital discharge) in a competing risks framework, where in-hospital mortality is considered to be a competing risk.[18] All study subjects were observed until time to in-hospital mortality or discharge, thus there was no censoring. There were a small number of patients who were discharged from the hospital, and later returned to the hospital and experienced in hospital mortality. The analysis treated the eventual in-hospital mortality for these patients as unobserved. Time to discharge and time to inpatient mortality are displayed by the nonparametric Aalen-Johansen curves. Inference is done via a Fine-Gray model for competing risks with robust variance estimation for the propensity-matched cohorts (http://github.com). In the Fine-Gray models, time to event is regressed on an indicator variable denoting treatment status with no other covariates.

## Results

In our primary intention to treat analysis, 188 treatment and control subjects were retained after propensity score matching for 376 total patients included in the trial (**Figure 1**). The variance ratio was used to assess covariate variance balance, with balance defined to be a variance ratio < 2.[19] Covariate balance was achieved after propensity score matching for the intention to treat analysis (**Figure 2**). Propensity score matching was also performed for the sub-analyses that limited the study population based on time to infusion. Covariate balance was achieved for the 7 or fewer days to infusion analysis and was nearly, but not achieved for the 3 or fewer days to infusion analysis (**Supplementary Figures 1&2**). Subject characteristics for the propensity score matched intention to treat analysis are displayed in **Table 1**. Overall, the cohorts enrolled represented the regional outbreak with higher proportion of older adult patients (mean 58.9 years), male patients (56.9%), and 50.8% of patients in the study identified as having a Hispanic ethnicity. In the total cohort, 67.2% of CCP-treated patients identified as white or Caucasian, 15.7% identified as >1 race, 10.4% identified as black or African American, and 4.8% identified as Asian. During the study, 38.3% of the patients were enrolled during May of 2020 and 26.3% of the patients were enrolled during July of 2020 (**Table 1**). Patients were matched based on month of enrollment due to the rapid changing treatment approaches throughout the pandemic. Comorbid medical diagnoses including hypertension, diabetes mellitus, and obesity were found in 42-60% of the patients throughout the study and were matched at baseline for each individual.

**Table 1:**
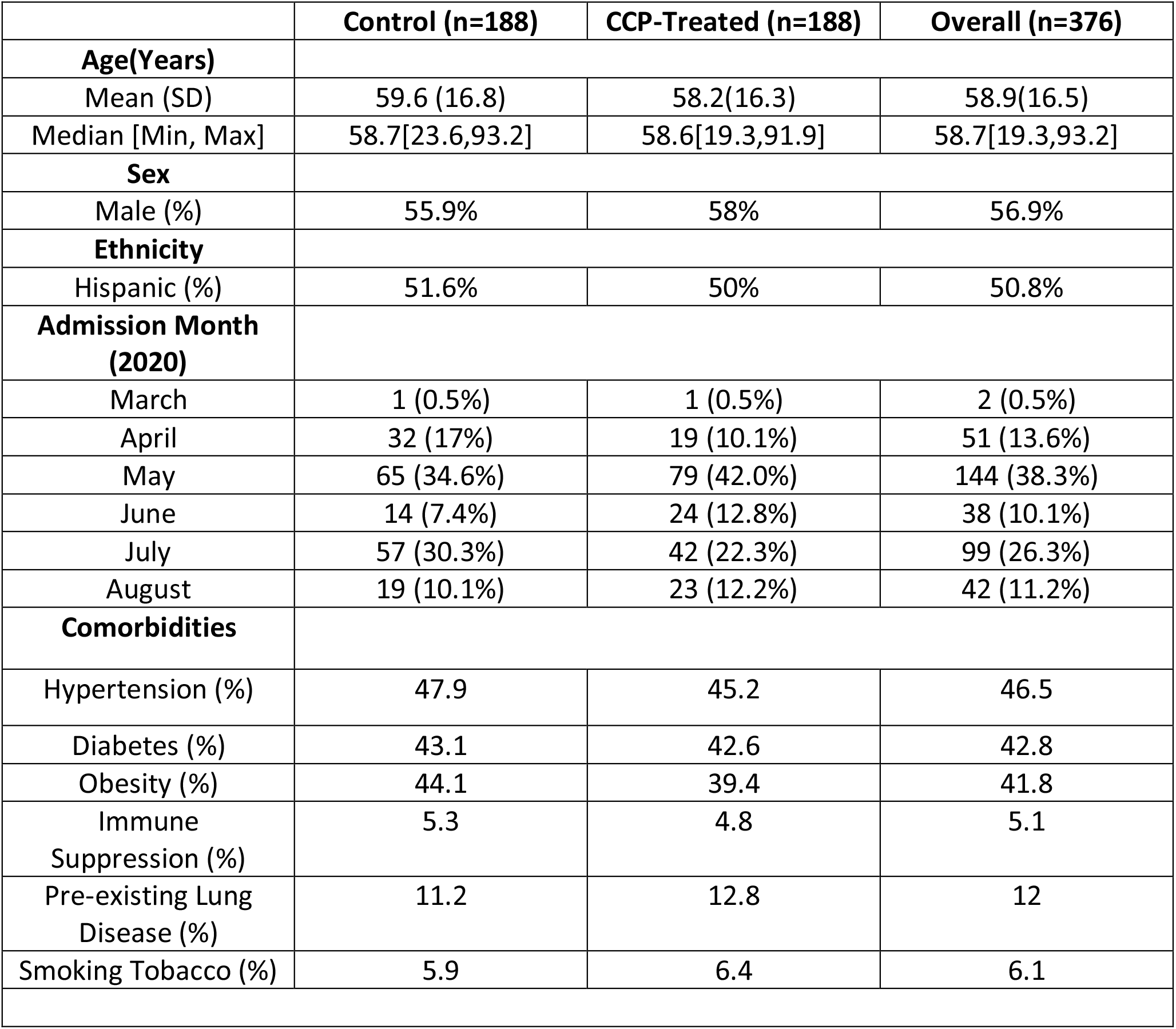
Demographic Data for Propensity score matched control and CCP-treated COVID-19 patients.

**Figure 1.**
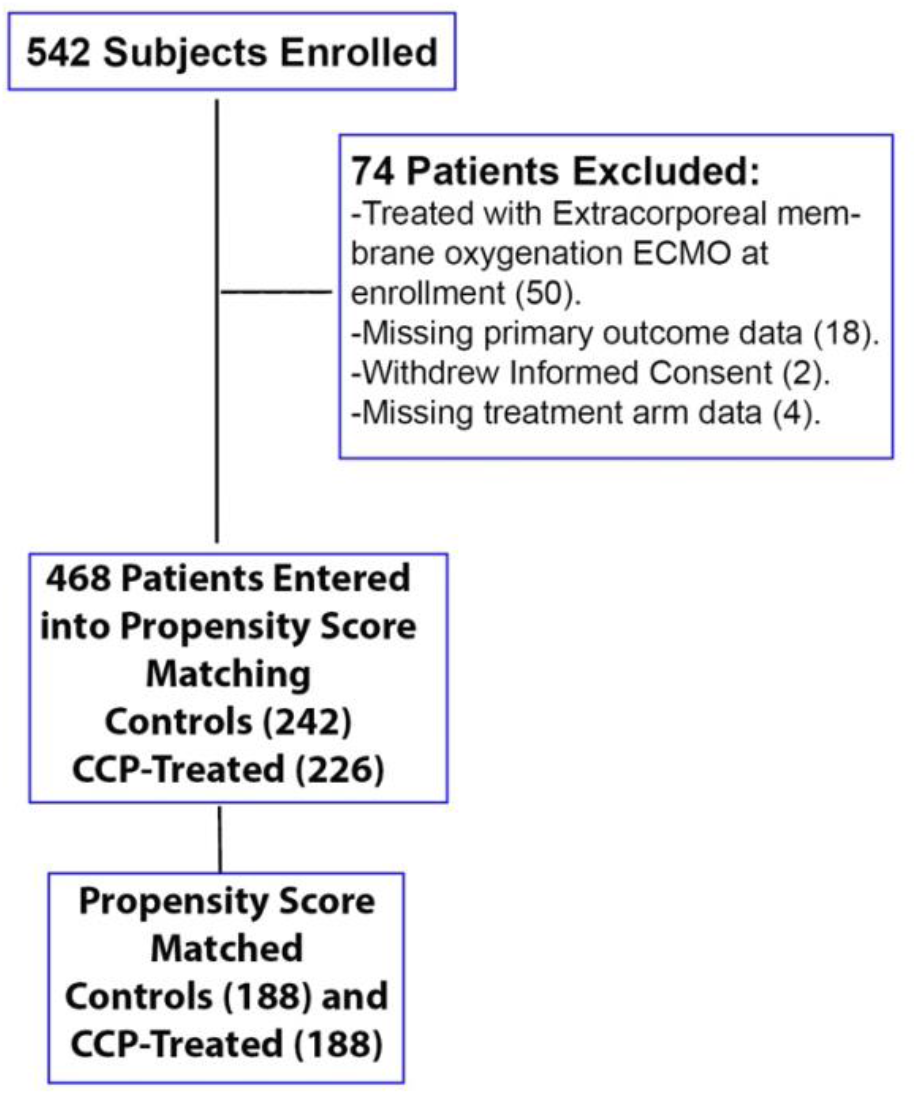
Flow-chart of patient enrollment and Propensity Score Matching.

**Figure 2.**
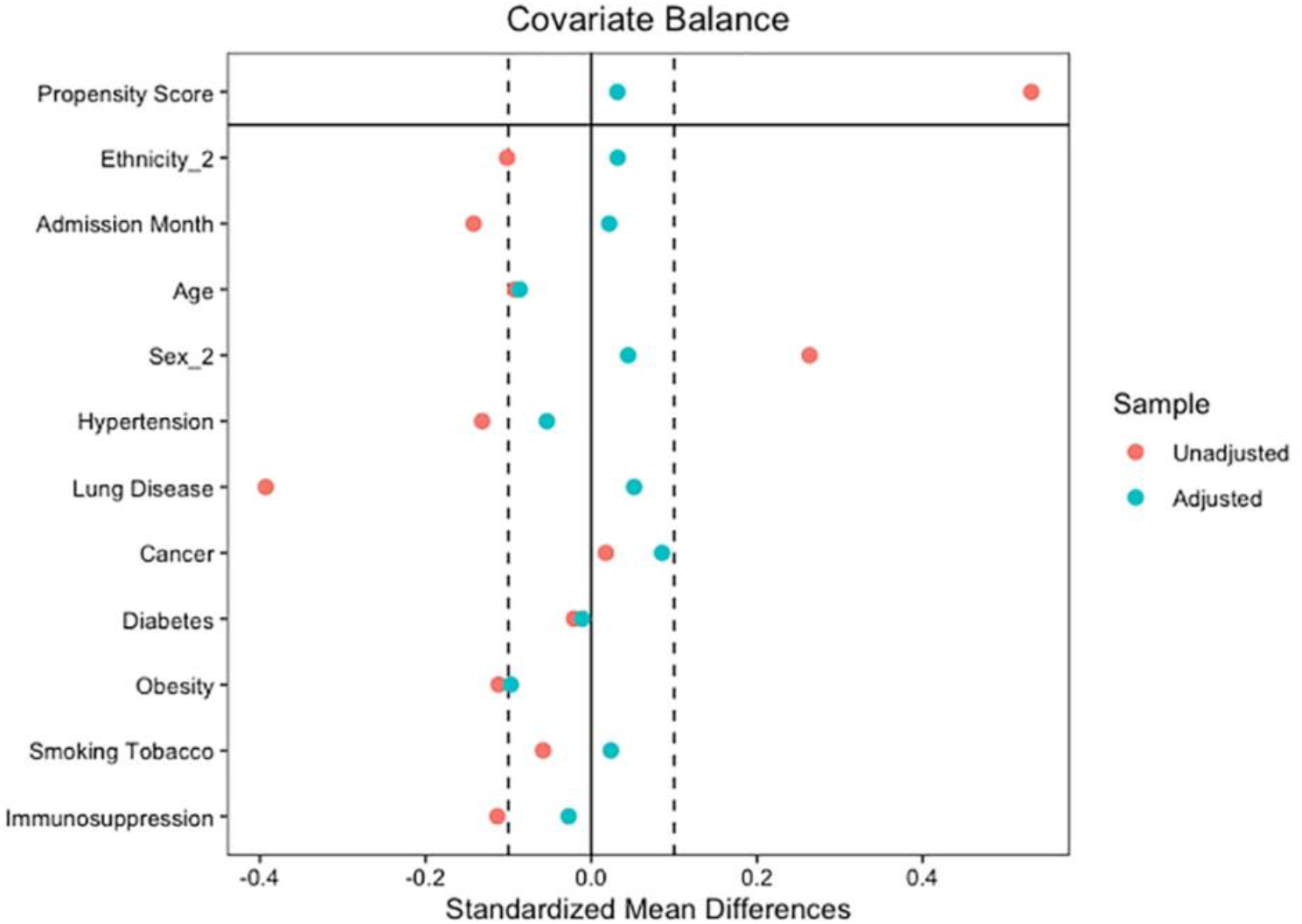
Love plot showing covariate balance in full data set analysis before (red) and after (green) propensity score matching.

A total of 468 subjects, 226 in the treatment arm and 242 in the control arm, underwent propensity score matching (**Figure 1**). For the intention to treat analysis, 376 one-to-one matched CCP treatment and control subjects remained after propensity score matching. Analysis of patients who received CCP treatment at 7 days or less of hospitalization resulted in 358 total subjects after matching. For analysis of patients who received CCP treatment at 3 or fewer days of hospitalization, 322 total subjects remained after matching.

Overall, treatment of hospitalized COVID19 patients with CCP was associated with a longer hospitalization calculated as a hazard ratio (0.67) for hospital discharge favoring control COVID19 patients compared to CCP-treated patients (p=0.00053, **Table 2**). As a secondary endpoint, there was no difference in inpatient mortality (p=0.47). Column 4 displays the estimated hazard ratio of treatment status (compared to control) on hospital discharge and inpatient mortality (**Table 2**). As these coefficients were estimated via Fine-Gray models, they should be interpreted as changes in the sub-distribution hazard function for each event type.[20] Values less than one indicate that treatment status reduces the hazard of the corresponding event. The estimated Aalen-Johansen incidence curves for hospital discharge and in-hospital mortality for each matched cohort show the probability of hospital discharge or inpatient mortality over time (**Figure 3**).

**Table 2:** Number of patients in the matched treatment and control arms that experience primary (hospital discharge) or secondary (inpatient mortality) outcomes.

| Propensity Matched Cohorts |  |  |  |  |
| --- | --- | --- | --- | --- |
| Outcome Measure | Control<br>(n=188) | CCP Treated<br>(n=188) | Hazard Ratio<br>(CI) | P-value |
| Hospital Discharge | 165 | 161 | 0.67(0.54,0.85) | 0.00053 |
| Inpatient Mortality | 23 | 27 | 0.81(0.46,1.43) | 0.47 |
| CCP Infusion within 7 days of Admission |  |  |  |  |
| Outcome Measure | Control<br>(n=179) | CCP Treated<br>(n=179) | Hazard Ratio | P-value |
| Hospital Discharge | 158 | 155 | 0.84 (0.67,1.04) | 0.11 |
| Inpatient Mortality | 21 | 24 | 1.08 (0.59,1.97) | 0.81 |
| CCP Infusion within 3 days of Admission |  |  |  |  |
| Outcome Measure | Control<br>(n=161) | CCP Treated<br>(n=161) | Hazard Ratio | P-value |
| Hospital Discharge | 139 | 141 | 0.77 (0.42,1.42) | 0.41 |
| Inpatient Mortality | 22 | 20 | 0.83 (0.66,1.06) | 0.13 |

CCP = COVID19 Convalescent plasma, CI=confidence interval

**Figure 3.**
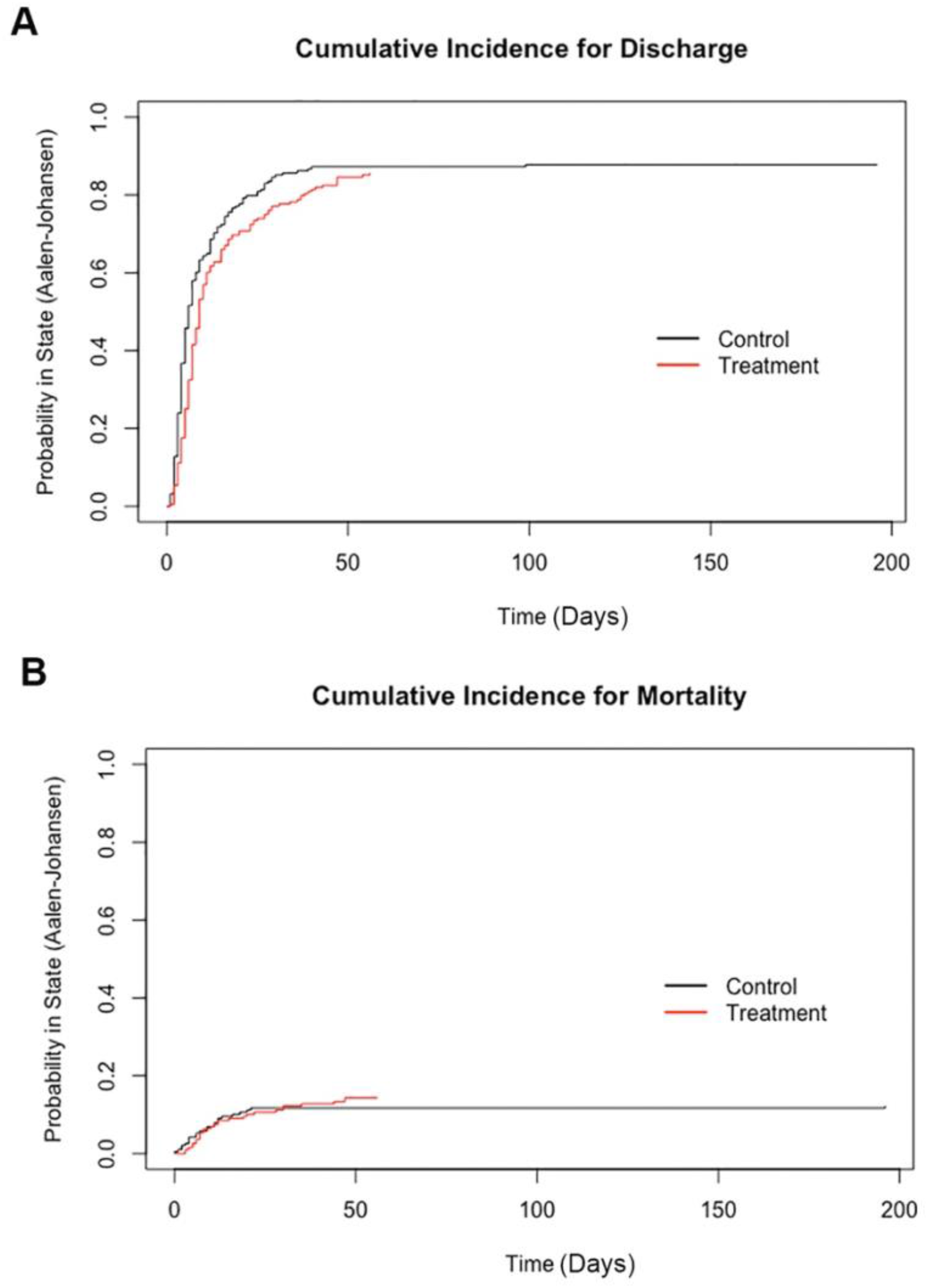
Cumulative incidence curves. **A)** Probability of hospital discharge over time in days. **B)** Probability of inpatient mortality over time in days. Estimated using Aalen-Johansen analysis.

Analysis of COVID19 patients that received CCP treatment within 7 or 3 days of admission revealed no significant difference in hospital discharge or inpatient mortality in CCP treated patients compared to control patients (**Table 2**). The Aalen-Johansen curves for CCP-treated patients within 7 or 3 days of admission also show no significant difference in hospital discharge or inpatient mortality when compared to control COVID19 patients (**Supplementary Figures 3-6**).

Four patients had documented adverse events associated with CCP infusion. Three patients had allergic reactions with pruritus, rash, or urticaria, which resolved following treatment with acetaminophen and antihistamines. One patient developed a febrile transfusion reaction which improved with acetaminophen therapy. The transfusion was stopped in the patient after 60mL, and the patient was included in the analysis.

## Discussion

In this prospective, open-label, multicenter cohort-controlled study, we found that treatment of hospitalized COVID19 patients with CCP did not significantly improve time to hospital discharge or inpatient mortality compared to propensity matched controls. Given that hospital discharge is considered a positive endpoint, the fact that treatment reduces the hazard of hospital discharge is evidence that CCP treatment may have a negative effect on the prognosis of study participants. However, this statistically significant effect disappears in the two sub-analyses. This may be because study participants who received convalescent plasma more than 7 days after hospital admission differ systematically from the rest of the study population. For example, the patients that received CCP after 7 days may bias the treatment group to more severe disease. We also found that CCP treatment status does not have a statistically significant effect on the hazard of inpatient mortality. It would be difficult to detect an effect of convalescent plasma treatment on the hazard of inpatient mortality in this study given the relatively few number of inpatient deaths.

Previous, mostly non-controlled trials have suggested a potential benefit of CCP therapy in hospitalized COVID19 patients. In a retrospective matched cohort study, patients receiving CCP had decreased 7-day and 14-day mortality but no statistical difference in 28-day mortality. Similar to our study, the length of hospital stay was increased in convalescent plasma group suggesting possible selection bias.[7] A propensity matched study performed at Houston Methodist hospital from March 2020 through September 2020 showed a significant decrease in mortality for patients transfused within 72 hours of admission with plasma containing anti-RBD IgG titer of > 1:1350. No mortality benefit was noted in patients who received RBD IgG titer of < 1: 1350 or were intubated at the time of admission.[6] In a cohort of 3082 patients who received convalescent plasma through the Mayo Clinic initiated expanded access protocol, non-mechanically ventilated patients who received high titer plasma had lower relative risk for death as compared to patients in low titer group. However, this study did not have a control group.[5]

Some randomized studies of CCP therapy in hospitalized COVID19 patients have shown no evidence of a clinical benefit. An open label RCT done earlier in the pandemic in China enrolled 103 patients and had to be terminated early due to decline in cases. This study did not show any benefit of convalescent plasma but was underpowered for the intended end-points.[21] A smaller study performed in Argentina of older adults who received convalescent plasma within 72 hours of symptom onset showed reduced risk of progression in patients receiving convalescent plasma.[22] In the PLACID trial, an open label multicenter RCT in India, 2 units of convalescent plasma was transfused. Neutralizing antibody was not found in 20% of the transfused plasma and median neutralizing antibody ranged widely from 1:30-1:240. In a smaller subgroup of patients with pre-existing neutralizing antibody, no benefit was noted[23].

The ConCOVID study was a randomized controlled trial comparing CCP with standard of care therapy. The study was discontinued early after enrollment of 86 patients (43 in each group) because a majority of enrolled patients had high titres of neutralizing antibody at the time of study enrollment and it was considered futile to continue further with the study. In this small subset of patients, no significant difference was noted in mortality or improvement in disease severity.[24]

Similar to prior studies, we found that convalescent plasma transfusion was well tolerated with rare adverse events. [22, 25, 26] In our study, we did not document any major transfusion reactions including transfusion-associated acute lung injury or hemolytic reactions. Minor transfusion reactions were noted in 4 of 239 patients (0.017%).

This study has several strengths including the multicenter design that included 16 academic and non-academic community hospitals throughout the Colorado front range region. This resulted in recruitment of hospitalized COVID19 patients that represented the demographics of the regional COVID19 pandemic with inclusion and analysis of a diverse cohort of patients. The size of the cohorts analyzed, and the matched control cohort were also strengths of the study. During this trial, there were no approved standard therapies including remdesivir or dexamethasone at the time of study. Weaknesses of the study include the open-label, non-randomized study design and lack of a placebo control group. While all CCP was evaluated for presence of SARS-CoV2 specific antibody, many of the units were retrospectively tested through individual regional plasma donation centers resulting in variability in the data. However, this approach also represented CCP treatment and distribution in the community during the pandemic.

## Conclusion

The risks of CCP-treatment in hospitalized COVID19 patients are minimal, this study shows that treatment with CCP for hospitalized COVID19 patients provides no significant improvement in length of hospitalization or inpatient mortality. Ongoing multicenter, randomized controlled trials of CCP treatment for hospitalized and outpatient COVID19 patients are critical to define a potential role for CCP therapy in the SARS-CoV-2 epidemic.

## Data Availability

All data related to the trial are available per University of Colorado data sharing and IRB guidelines.

## Acknowledgements

This study was supported in part by grant provided through Colorado Clinical and Translational Sciences Institute (CTSA Grant UL1 TR002535) to JDB. The study was also supported by institutional funds from University of Colorado Anschutz Medical Campus, Department of Medicine to JDB.

---

**Supplementary Figure 1.**
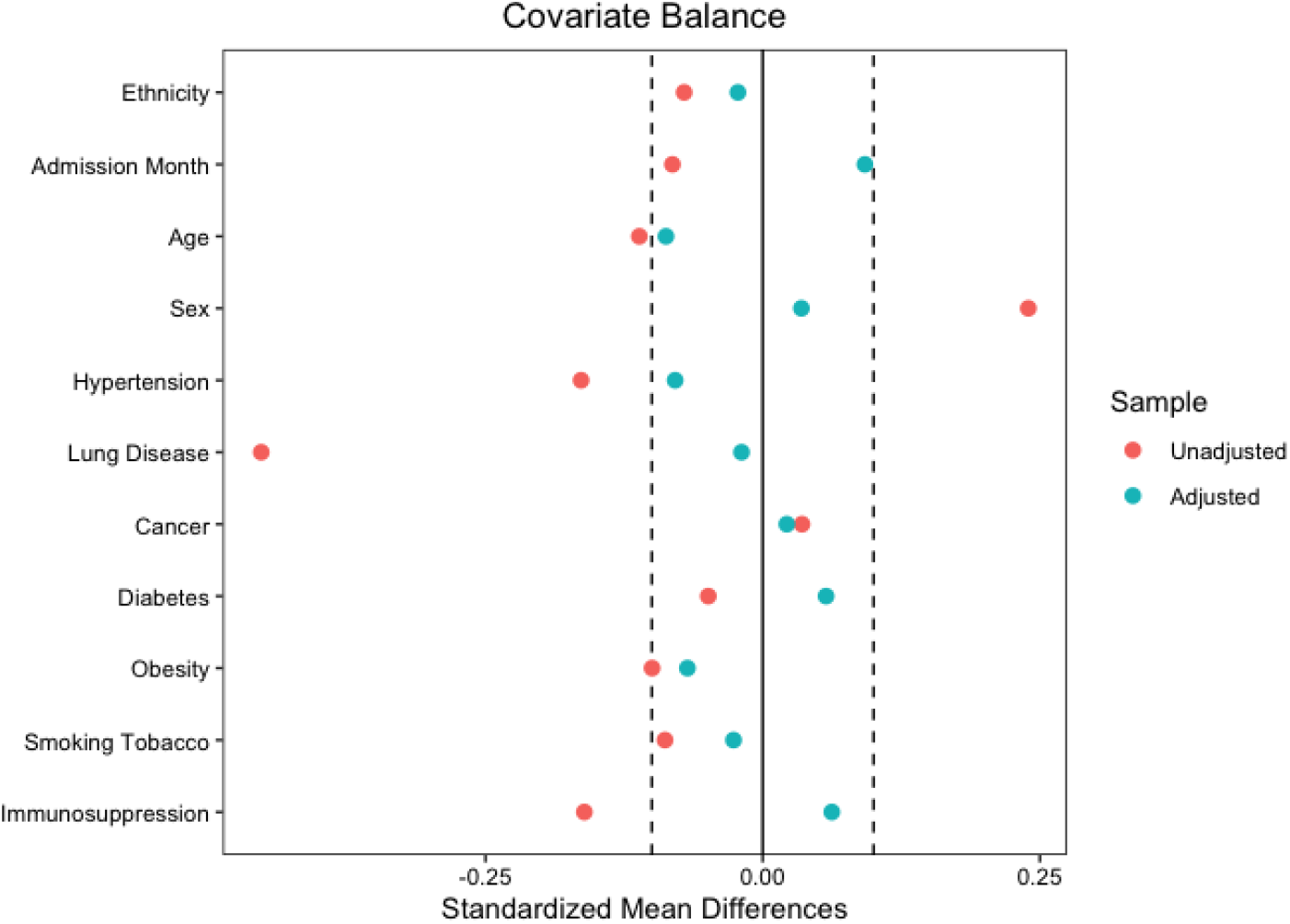
Love plot showing covariate balance in the 7 or fewer days to infusion analysis before and after propensity score matching.

**Supplementary Figure 2.**
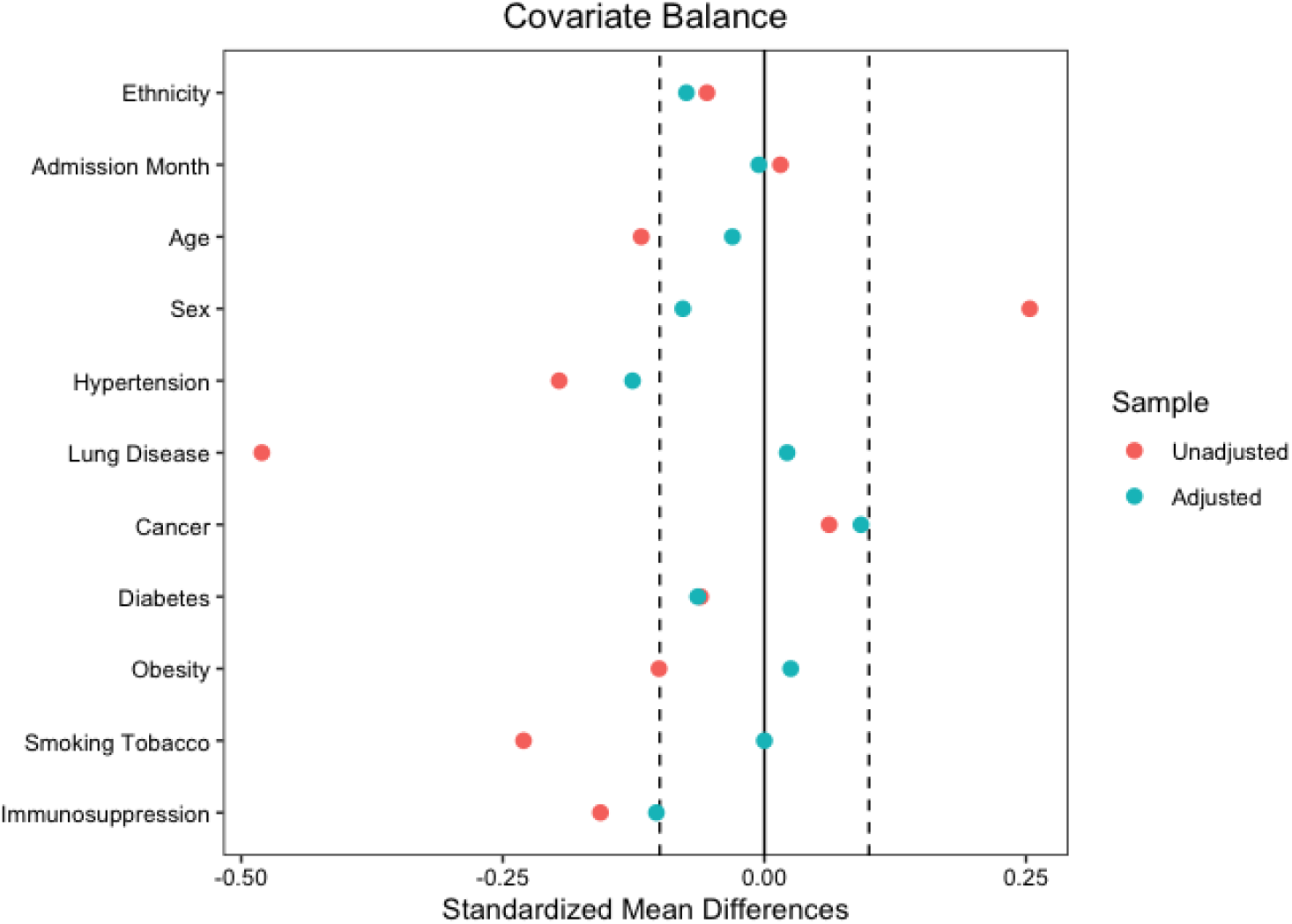
Love plot showing covariate balance in the 3 or fewer days to infusion analysis before and after propensity score matching.

**Supplementary Figure 3.**
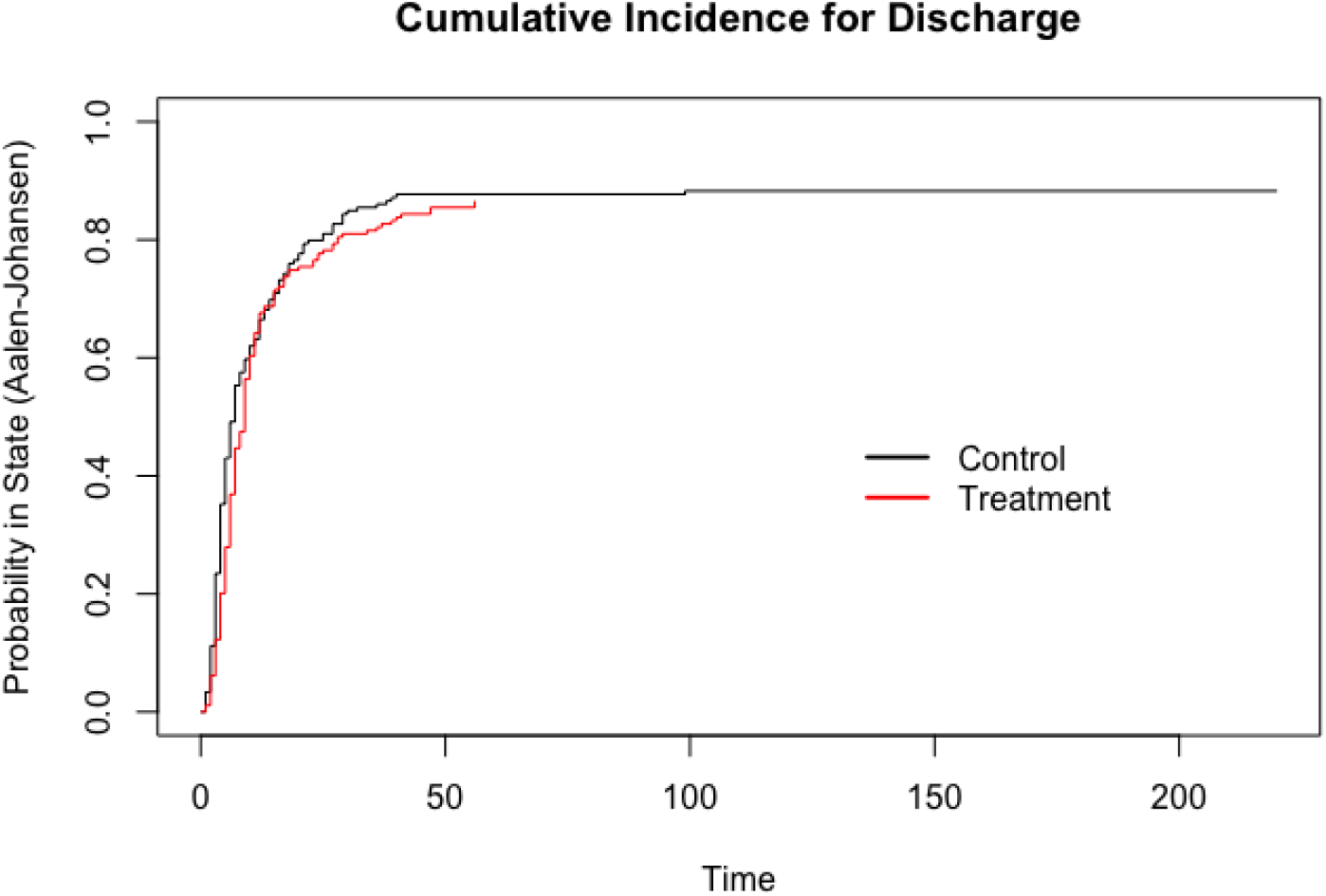
Cumulative incidence curve for hospital discharge in the 7 or fewer days to infusion analysis. Estimated via Aalen-Johansen.

**Supplementary Figure 4.**
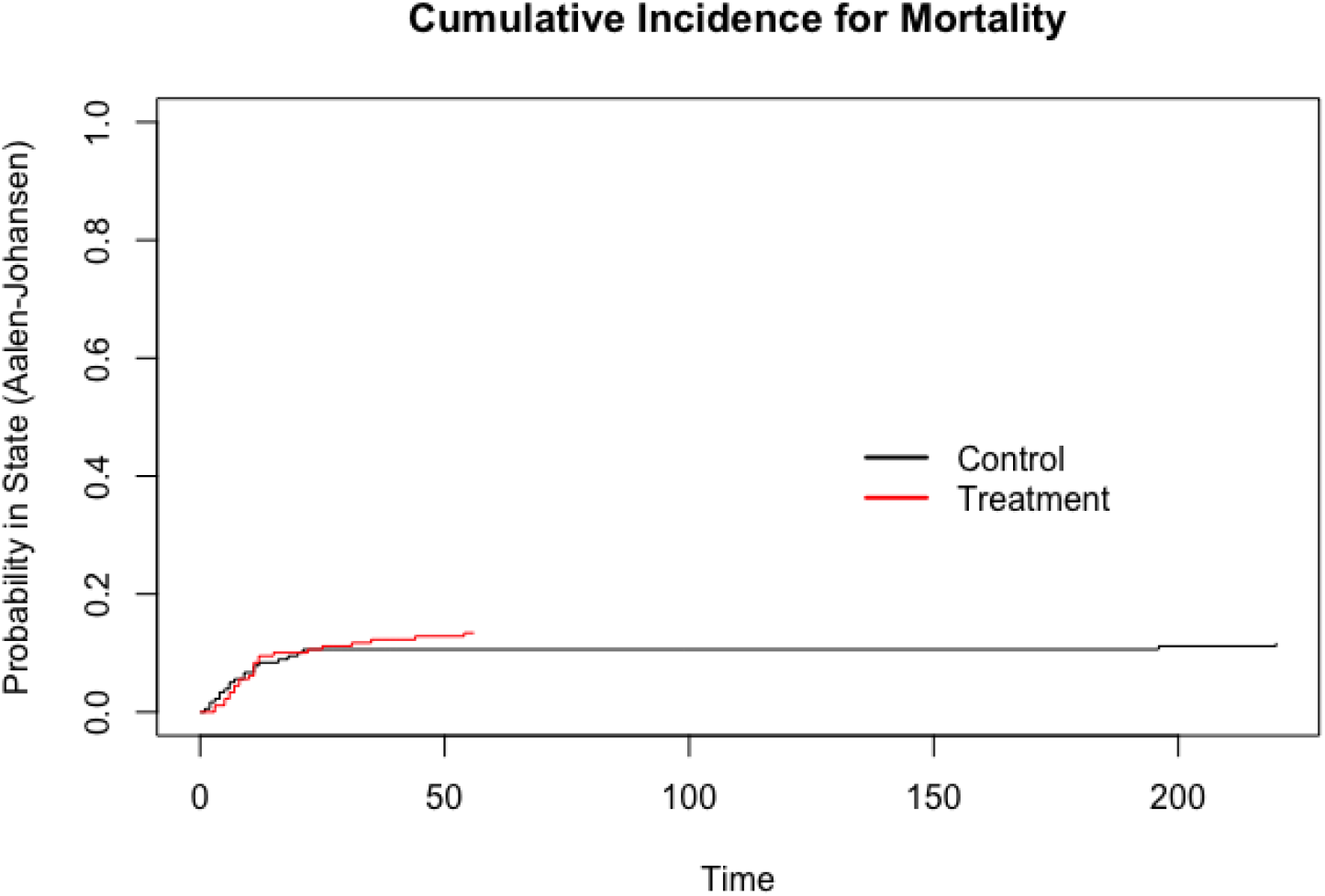
Cumulative incidence curve for mortality in the 7 or fewer days to infusion analysis. Estimated via Aalen-Johansen.

**Supplementary Figure 5.**
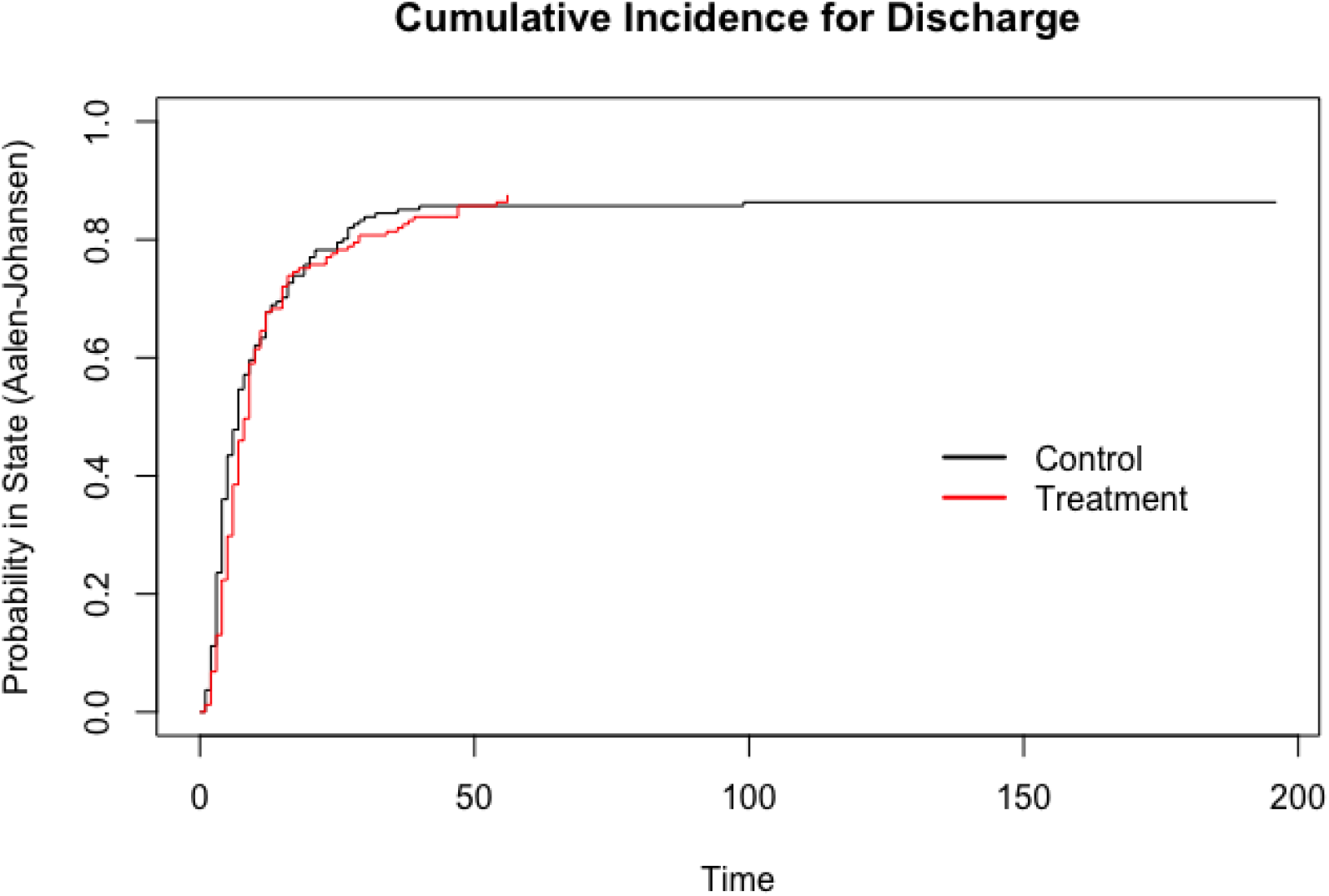
Cumulative incidence curve for hospital discharge in the 3 or fewer days to infusion analysis. Estimated via Aalen-Johansen.

**Supplementary Figure 6.**
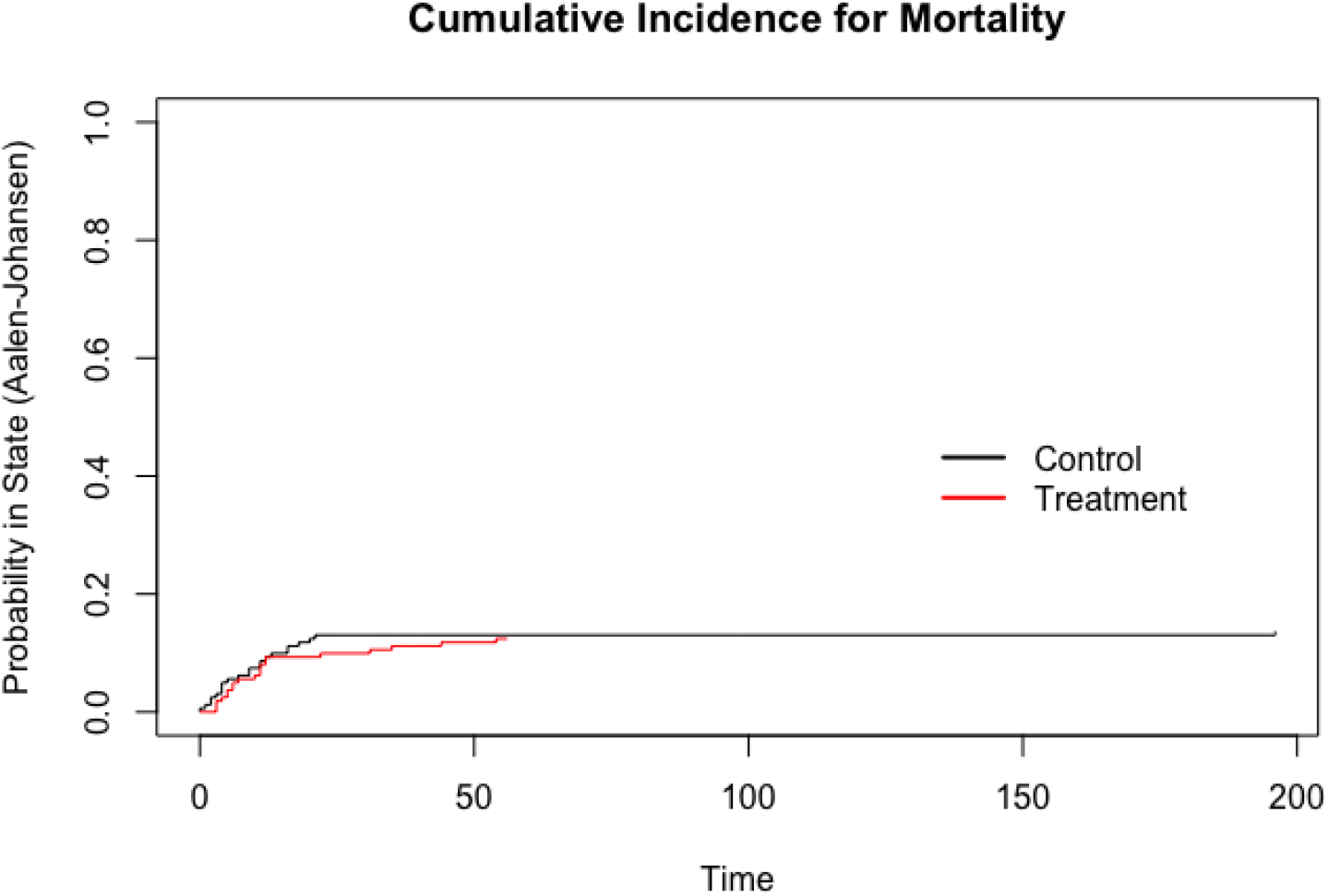
Cumulative incidence curve for mortality in the 3 or fewer days to infusion analysis. Estimated via Aalen-Johansen.

